# Changing awareness and sources of tobacco and e-cigarettes among children in Great Britain

**DOI:** 10.1101/2023.05.11.23289841

**Authors:** Jennie C. Parnham, Charlotte Vrinten, Hazel Cheeseman, Laura Bunce, Nicholas S Hopkinson, Filippos T Filippidis, Anthony A. Laverty

## Abstract

**Introduction:** It is illegal in the UK to sell tobacco or nicotine e-cigarettes to people under the age of 18 years, as is displaying tobacco cigarettes at the point of sale. This paper examined changes in exposure to display of these products in shops and sources of these products among adolescent users over time.

**Methods:** Data from representative repeated online cross-sectional surveys of youth in Great Britain (11-18 years) were used (2018-2022; n=12,445). Outcome measures included noticing product displays and sources of e-cigarettes and tobacco cigarettes. Logistic regressions examined the associations of these outcome variables over time and with sociodemographic variables.

**Results:** Of 12,040 participants with complete data, 10.1% used some form of nicotine product (4.2% cigarettes, 2.9% e-cigarettes, 3.0% both) at least occasionally. The likelihood of noticing tobacco cigarettes on display fell over time for both supermarkets (2018: 67.1% to 2022: 58.5%) and small shops (2018: 81.3% to 2022: 66.3%), but the likelihood of noticing e-cigarettes in supermarkets rose (2018: 57.4% to 2022: 66.5%). Sources of tobacco cigarettes did not differ over time, but e-cigarette users were more likely to get their e-cigarettes from small shops in 2022 (51.2%) vs. 2019 (34.2%) (Odds ratio 2.02; 95%CI 1.24, 3.29).

**Conclusion:** This study provides evidence that current policies to limit awareness of and access to both tobacco and e-cigarettes among adolescents in the UK may not be effective. UK policy on the advertising, promotion and sale of both tobacco and e-cigarettes need to be reinforced to deter use among children.

## INTRODUCTION

Advertising, promotion, and sponsorship of tobacco products are known to be key methods of encouraging tobacco use [1]. Children and young people are particular targets for such advertising, due to the need for the tobacco industry to recruit tobacco users, and increased susceptibility to pro-smoking messages [2]. The retail environment is a key location for such advertising and promotion, with evidence linking exposure to these to increased smoking desire and cigarette purchasing [3–5]. The World Health Organization (WHO) advocates a complete ban on the display of tobacco at the point of sale[6], although only 22% of the global population live in countries with total tobacco advertising bans[7]. Although there is less data on the issue of advertising and promotion of e-cigarette due to their recent emergence, the WHO recommends that member states regulate these products as appropriate to their circumstances [8].

Another key strategy to reduce uptake and use of tobacco among young people is reducing access through enforcing a minimum age of sale[9]. Increased minimum age of sale policies are designed to reduce direct access to tobacco products among children, as well as reducing access through friends and social sources [10]. These policies are known to be effective: for example, the rise in the legal age of sale from 16 to 18 years in England was linked to both an immediate and long-term fall in prevalence among 16 and 17 year olds[11,12]. For e-cigarettes the WHO also recommends minimum ages of sale for e-cigarettes[7]. The latest WHO report on the global tobacco epidemic found that 69 countries (40%) had minimum age of sale legislation for e-cigarettes, with 40 of these countries being in the European Region. This is compared to 90% of countries which have age-restriction for policies for tobacco.

The UK has legislation in place to restrict promotion, advertising, and sponsorship as well as access to tobacco products among children[13]. Tobacco displays at the point of sale were banned in two stages: in large shops (with >280⍰m^2^ floor area) in April 2012 and then in all shops from April 2015[14]. This legislation has been found to reduce exposure to tobacco advertising among children in both England and Scotland, although exposure levels have remained high[15,16]. Regulations for the advertising of e-cigarettes are less stringent, although this is not allowed on the TV, radio, online or in print media. [17], but there is no equivalent point of sale display ban for e-cigarettes in the UK. The sale of tobacco to people under the age of 18 is not allowed, nor is sale of e-cigarettes which contain nicotine. Despite these regulations, e-cigarette use is reported by approximately nine percent of 11-18 year olds (including occasional and regular use) with use more common among those reporting seeing e-cigarette promotion [18,19]. With rapid changes in the tobacco and e-cigarette market in recent years, including possible impacts of COVID-19, better data on levels of exposure to tobacco and e-cigarette advertising as well as where children are obtaining these products is needed. This paper examined changes in exposure to display of both tobacco and e-cigarettes among adolescents in Great Britain from 2018 to 2022, and examined the sources of tobacco and e-cigarettes among adolescent users.

## METHODS

### Data source

Data were taken from an annual cross-sectional survey of adolescents (11-18 years) the Action on Smoking and Health (ASH) Smokefree GB Youth Survey, 2018-2022. This online survey has been run by YouGov since 2013, in which a random sample of parents of 11–18-year-olds from their membership panel were invited to participate by email each year. Informed consent from both the parent and the child was given before the adolescent completed the questionnaire. The adolescent survey includes questions on their sociodemographic characteristics, their awareness of and attitudes towards tobacco and e-cigarettes product use, as well as their own use of these products. A total of 12,445 participants responded, of which 405 were excluded due to missing data (social grade n=164; product use n=241). Weights, supplied by the survey company, were used to ensure the responses were representative of all adolescents in Great Britain aged 11-18 years.

### Product-use

All participants were asked whether they used cigarettes and e-cigarettes, respectively. Participants were coded as ‘*Current cigarette users*’ if they responded ‘sometimes but less than one a week’, ‘between one and six cigarettes a week’ or ‘more than six cigarettes a week. Participants were coded as ‘*Former cigarette users*’ if they responded, ‘used in the past but not now’ or ‘tried once or twice’. Similarly, participants were coded as ‘*Current e-cigarette users*’ if they responded they use e-cigarettes ‘sometimes but no more than once a month’, ‘more than once a month, but less than once a week’, ‘more than once a week but not daily’ and ‘every day’. Participants were coded as ‘*Former e-cigarette users*’ if they responded, ‘used in the past but not now’ or ‘tried once or twice’. ‘*Non-users*’ were participants who responded that they had never used the product, for cigarettes and e-cigarettes respectively. A nicotine-product use summary variable combining answers to both e-cigarette use and tobacco smoking was also created. Participants were defined as a ‘*non-user*’ if they did not currently use tobacco cigarettes and e-cigarettes, an ‘*E-cigarette user only*’ if they reported currently using e-cigarettes but not tobacco cigarettes, a ‘*Tobacco cigarette user only*’ if they reported currently using tobacco cigarettes but not e-cigarettes and a ‘*Dual user*’ if they reported currently using tobacco cigarettes and e-cigarettes.

### Display of products

Participants who had responded they were aware of e-cigarettes (n=11,188) were asked “When you go into supermarkets, how often, if at all do you notice e-cigarettes on display?” with a similar question for small shops. The same two questions were asked about tobacco cigarettes in supermarkets and small shops. The six possible responses were recoded into a binary variable (‘*Notice*’ = ‘Every time’, ‘Most times’, ‘Sometimes’ and ‘Hardly ever’; ‘*Do not notice*’ = ‘Never’ and ‘I never go to supermarkets/small stores’).

### Sources of nicotine-products

Participants who currently used tobacco cigarettes or e-cigarettes were asked where they usually source these, with two separate questions asked to e-cigarette (n=717) and tobacco cigarette users (n=867). The participants could choose multiple sources from 16 categorical options and were given an option of an open-ended ‘other’ response. All of the responses were coded into four binary variables which were not mutually exclusive: ‘*Bought; supermarket*’, ‘*Bought: small shop*’ (such as newsagents, garages and vape shops), ‘*Bought: online*’, and ‘*Acquired other*’. The sources were categorised to distinguish areas which tobacco control policies directly affect access, such as age-restriction measures. Therefore, ‘*Acquired other*’ represents a range of sources, including acquiring from a friend or family member, or purchase from a street market. Only four individuals responded that they bought tobacco cigarettes online, therefore this was not used as a separate category in the analysis. See **Supplementary table 1** for a full list of responses and categorisation.

### Covariates

Covariates considered in the analysis included survey year (2018-2022), age group (11-13, 14-15, 16-17 and 18 years old), gender (male or female), social class (based on National Readership Survey (NRS) classification of occupations[20] and classified as ABC1 (higher) vs. C2DE (lower)), country (England, Scotland, or Wales), current e-cigarette use by others in the household (yes or no) and current tobacco smoking by others in the household (yes or no).

### Statistical analysis

Data collection on noticing display of tobacco and e-cigarette products began in 2018, while the question on sources of e-cigarettes was added in 2019. Weighted χ^2^ tests were used to determine difference in covariates and nicotine-product use across survey years.

To determine factors associated with noticing nicotine products on display in supermarkets and small shops, we ran logistic regression models separately for both e-cigarettes and tobacco cigarettes. Covariates included survey year, gender, age, social grade, country, current nicotine-product use status and e-cigarette and tobacco smoking in the household.

Logistic regression models were also used to examine factors associated with sources of nicotine products among current users. Separate models were run for e-cigarette and tobacco cigarette outcomes. Interactions were tested between age and survey year (as numerical variables) for each model.

All statistical analysis were weighted to ensure the findings were representative to adolescents in Great Britain aged 11-18 years.

### Sensitivity analysis

To test the results were robust to possible misclassification bias, the variables for noticing e-cigarettes and tobacco cigarettes in supermarket and small shops were recoded. The ‘*Do not notice*’ was recategorized to include ‘Hardly ever’, ‘Never’ and ‘I never go to supermarkets/small stores’. The logistic regression models were repeated with the recoded variables and shown in the supplementary files.

## RESULTS

There were 12,040 participants in the analytic sample, of which 10,453 had complete data on questions related to noticing nicotine products on display, 608 were current e-cigarette users with complete data on questions relating to source of e-cigarettes (years 2019-2022) and 831 were current tobacco cigarette users with complete data on questions relating to sources of tobacco cigarettes (years 2018-2022) (See **Supplementary Figure 1** for full details of exclusions).

In the full sample, 51% of participants were female, 71% were in higher (ABC1) social classes and 86% lived in England (**Table 1**). Across survey years, there were significant differences in the age distribution of participants, with a lower proportion of participants aged 18 years in 2021 compared with other yearsA greater proportion of participants were current e-cigarette (5.9%) or dual users (4.3%) in 2022 compared to 2018 (both 1.9% in 2018). The proportion of participants living in a household where e-cigarettes were used was higher in 2022 (21.3%) compared with 2018 (17.4%), whereas for smoking these proportions were similar (8.5% in 2022 compared with 9.0% in 2018).

**Table 1.**
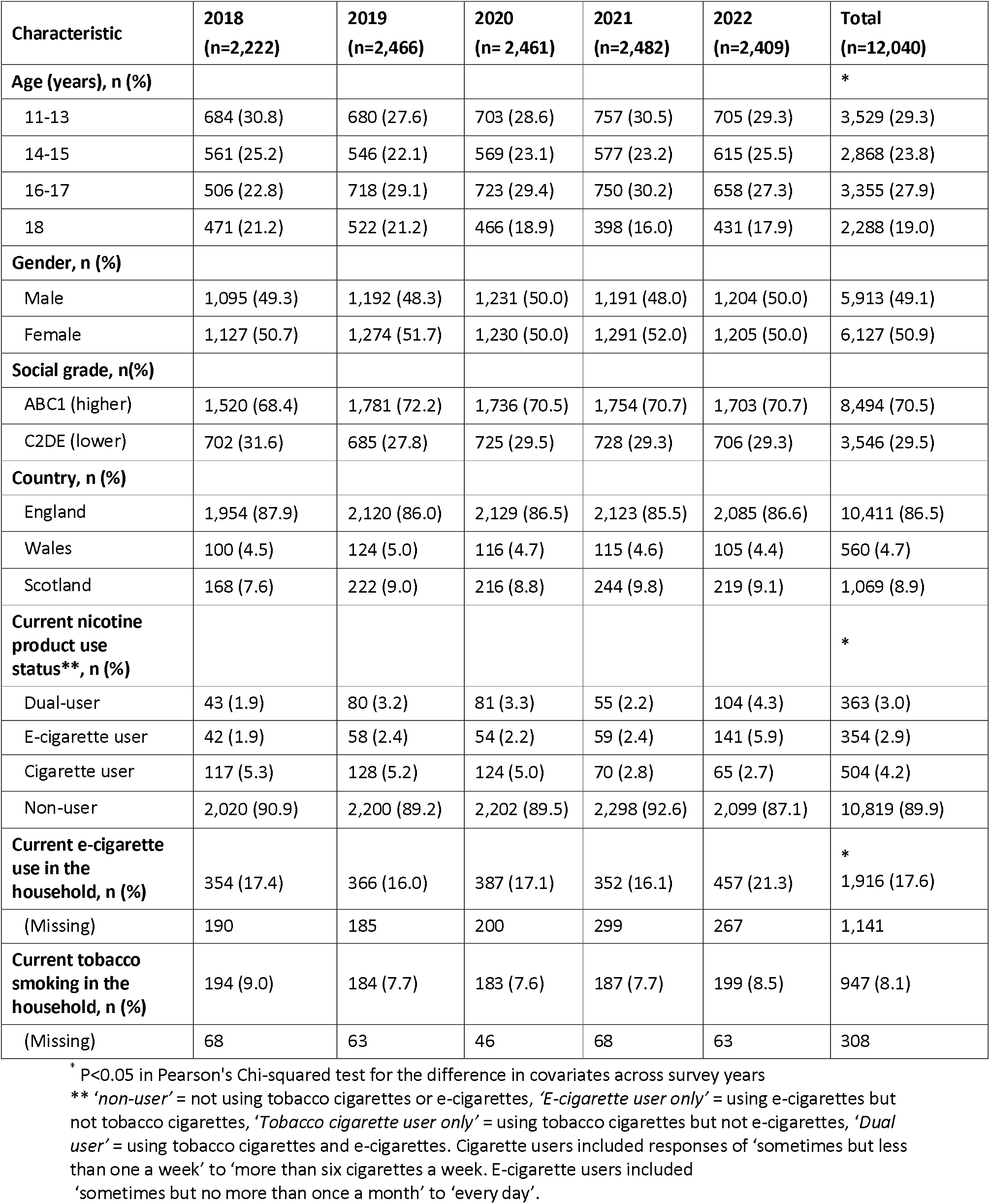
Characteristics of 12,040 UK adolescents who participated in the ASH Smokefree GB survey 2018-2022

### Factors associated with noticing nicotine-products on display in supermarkets and small stores

The proportion of the sample that noticed tobacco cigarettes on sale in supermarkets fell from 67% in 2018 to 59% in 2022 (**Table 2, Model 1**). In the adjusted logistic regression, this corresponded to a 29% (OR 0.71, 95% CI 0.61, 0.82) lower odds of noticing tobacco cigarettes on display in supermarkets in 2022 than 2018. Noticing cigarettes in small shops also fell over time (2018: 81%; 2022: 66%). There was a lower likelihood of noticing cigarettes in small shops for all years (2019-2022) compared to 2018.

**Table 2.**
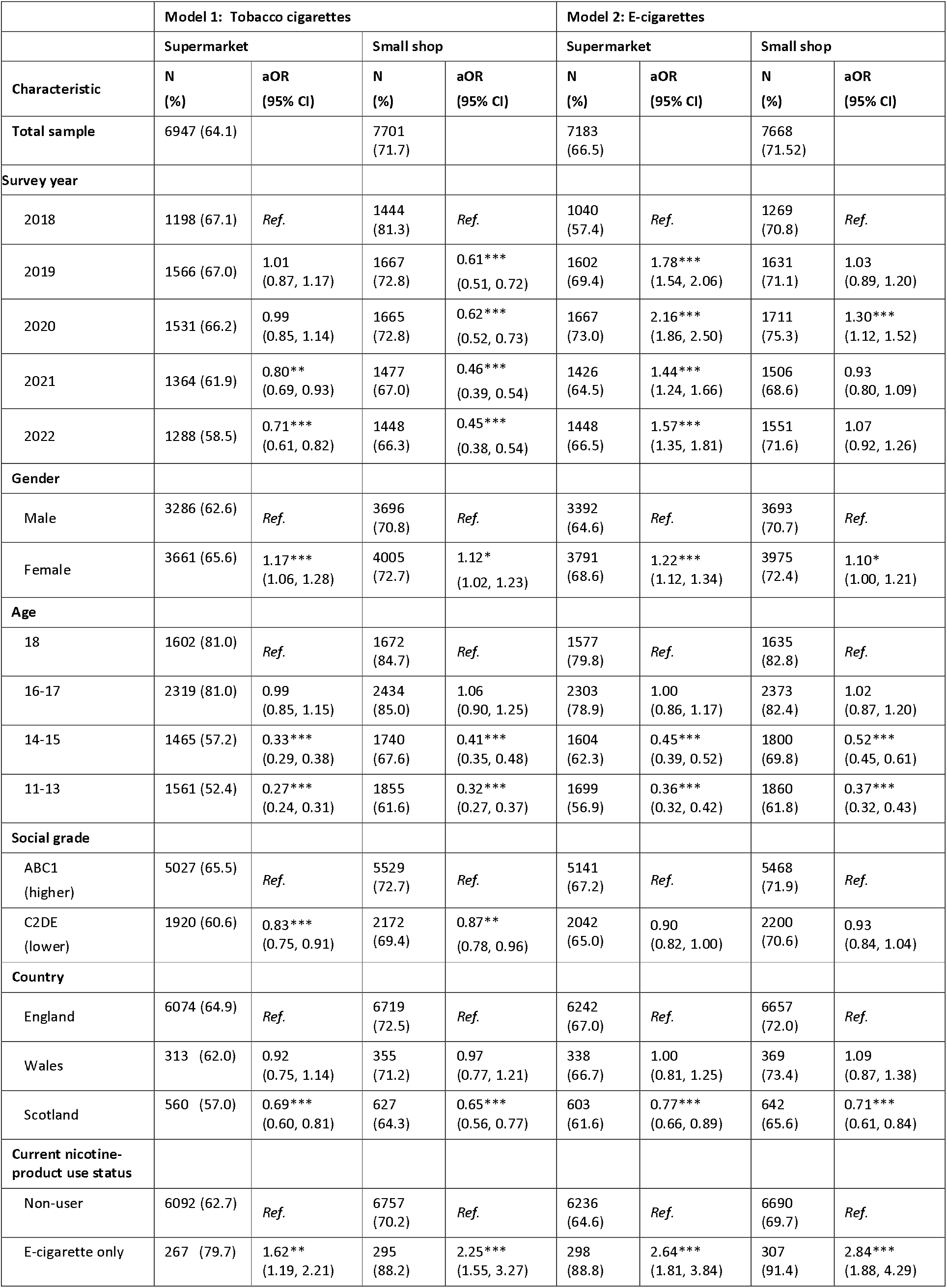
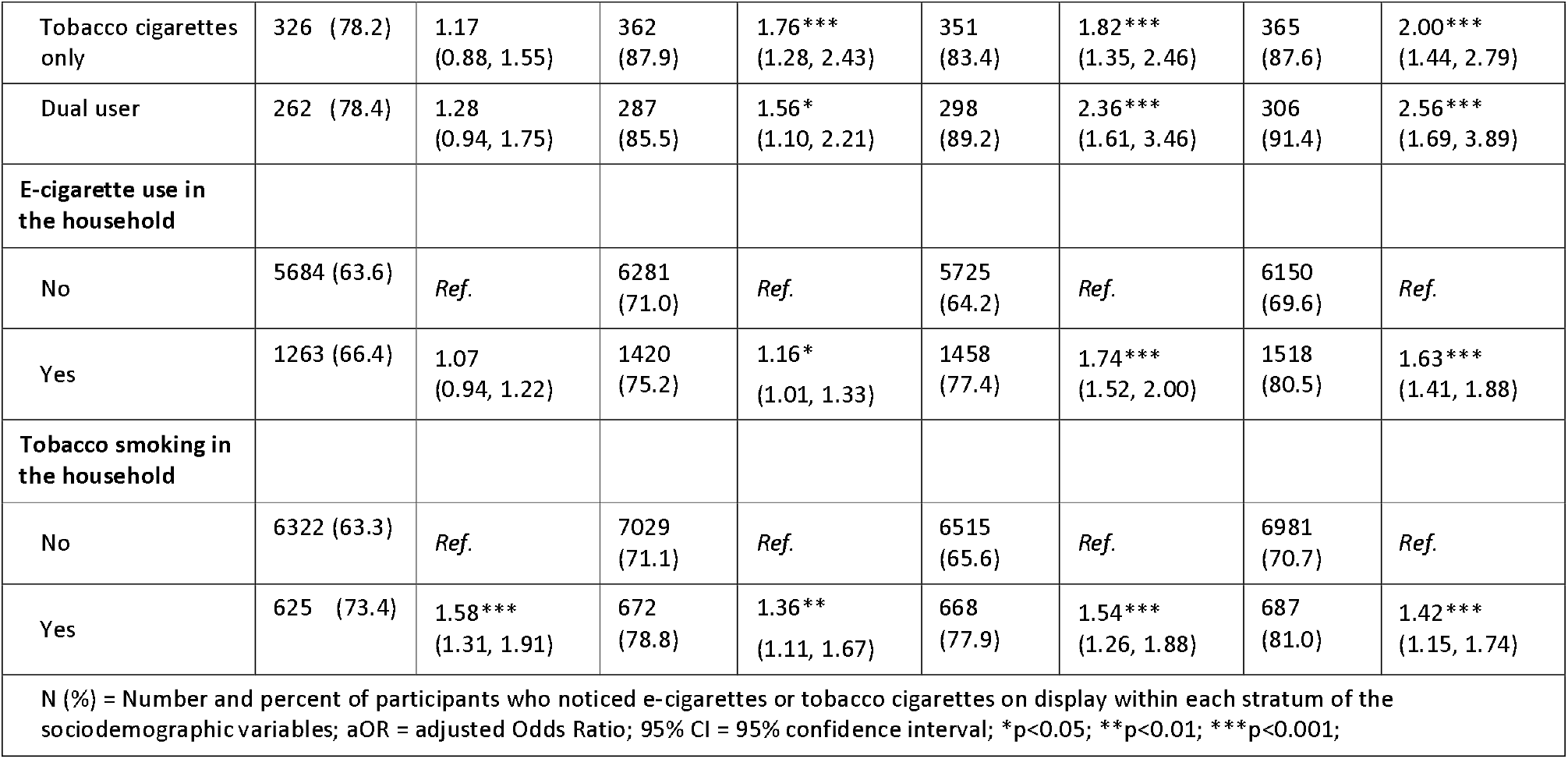
Adjusted logistic regression showing the likelihood of noticing e-cigarettes and tobacco cigarettes on sale in supermarkets and small shops (n=10,453).

Noticing tobacco cigarettes on display in supermarkets and small shops was significantly less likely for those who were younger (11-13 and 14-15, vs 18 years), those in lower compared to higher social classes and Scottish compared to English participants. Those who lived in a household with current tobacco cigarette use were significantly lower more likely to notice tobacco cigarettes in supermarkets and small shops. All forms of nicotine product use (e-cigarette only, tobacco cigarette only and dual use) were associated with a significantly lower higher likelihood of noticing tobacco cigarettes in small shops compared to non-users, but in supermarkets, only being an e-cigarette user was associated with a higher likelihood when compared to non-users.

The proportion of the sample that noticed e-cigarettes in supermarkets rose from 57.4% in 2018 to 66.5% in 2022 (**Table 2, Model 2**). In the adjusted logistic regression, there was a higher odds of noticing e-cigarettes in supermarkets for all years (2019-2022) than in 2018. Participants were more likely to notice tobacco cigarettes in small shops than supermarkets at baseline (81.3% vs. 67.1%), with the same being true for e-cigarettes. The proportion of participants noticing e-cigarettes in small shops was higher at baseline than for supermarkets (2018 e-cigarettes: 70.8% vs 2018 tobacco cigarettes: 57.4%). Overall there was not strong evidence that the likelihood of noticing e-cigarettes in small shops changed over time.

The likelihood of noticing e-cigarettes on sale in both supermarkets and small shops was statistically significantly lower for younger compared to older participants (11-13 and 14-15, vs 18 years) and those in Scotland compared to England. The likelihood was significantly lower higher for females compared to males, those who used nicotine products (e-cigarette only, tobacco cigarette only and dual use) compared with non-users, and for participants who lived in households where people used e-cigarettes and smoked tobacco cigarettes compared with those who did not.

#### Sources of tobacco cigarettes among current tobacco cigarette users

Overall, 25.1% of participants who smoked cigarettes sourced them from supermarkets, 50.6% sourced them from small shops and 58.1% acquired them from other sources (**Table 3**). In adjusted logistic regression, there was no evidence that sources of tobacco cigarettes changed over time. Compared to 18-year-olds users, younger cigarette users (16-17 and 14-15 years) were less likely to buy their cigarettes from both supermarkets and small shops. However, 16-17- and 14–15-year-olds were roughly three times more likely to acquire tobacco cigarettes from other sources than those aged 18 years (OR 3.51; 95% CI 2.11, 5.85) and OR 2.81; 95% CI 2.02, 3,91, respectively). Participants living in a household with someone smoking tobacco cigarettes were more likely to buy their tobacco cigarettes from a supermarket (OR 1.61; 95% CI 1.01, 2.59) and small shops (OR 2.26; 95% CI 1.46, 3.50).

**Table 3.**
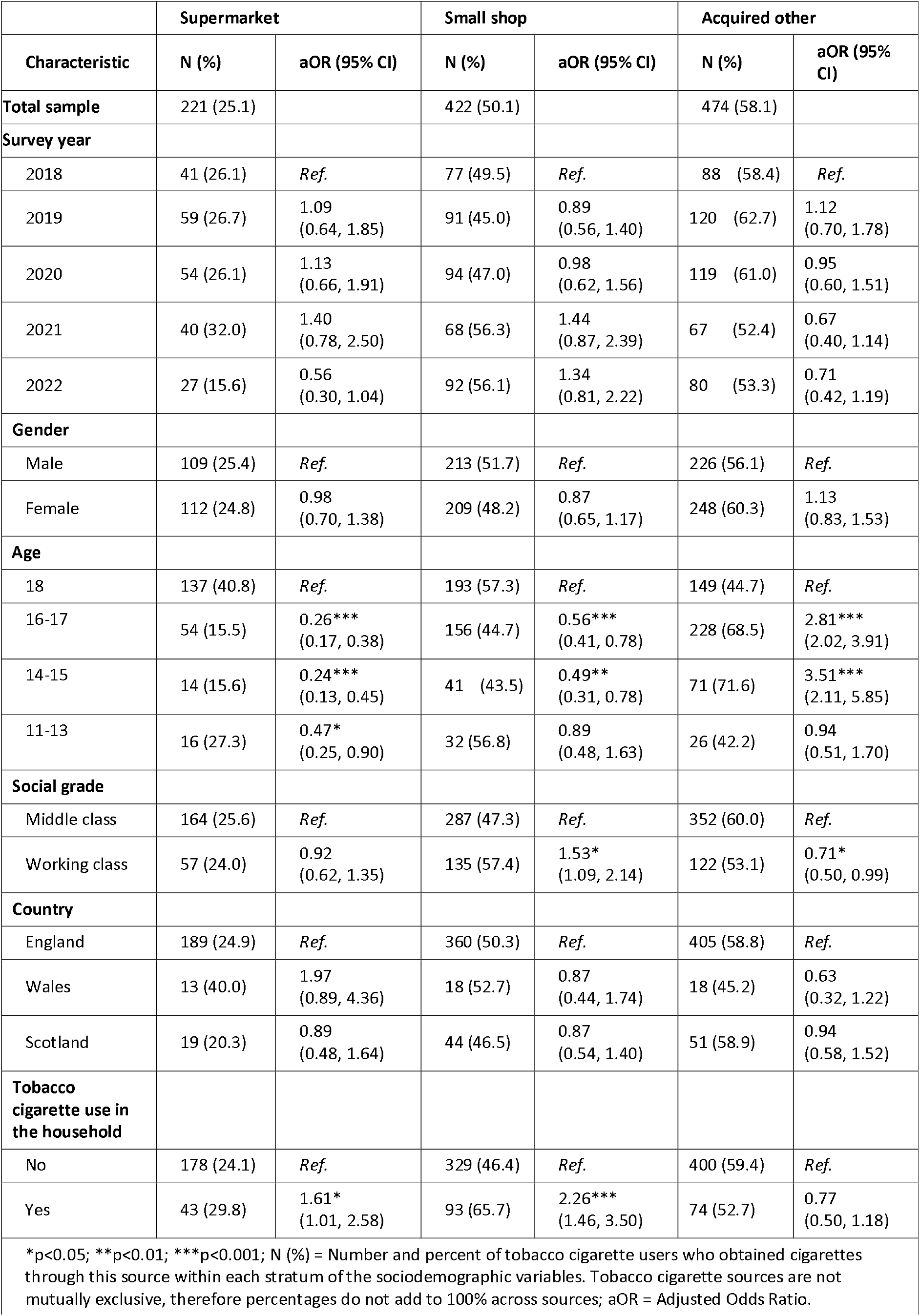
Fully adjusted and weighted logistic regression for likelihood of acquiring tobacco cigarettes from different sources in UK adolescents (n=831)

Interactions between age and survey year were tested for all regression models and were statistically significant for buying tobacco cigarettes from supermarkets (**Figure 1.A**). In the younger participants, there was a significant decreasing trend in buying cigarettes from supermarkets from over 30% in 2018 to less than 10% in 2022. The likelihood had a slight increase for the older participants, but the confidence intervals between 2018-2022 overlapped.

**Figure 1.**
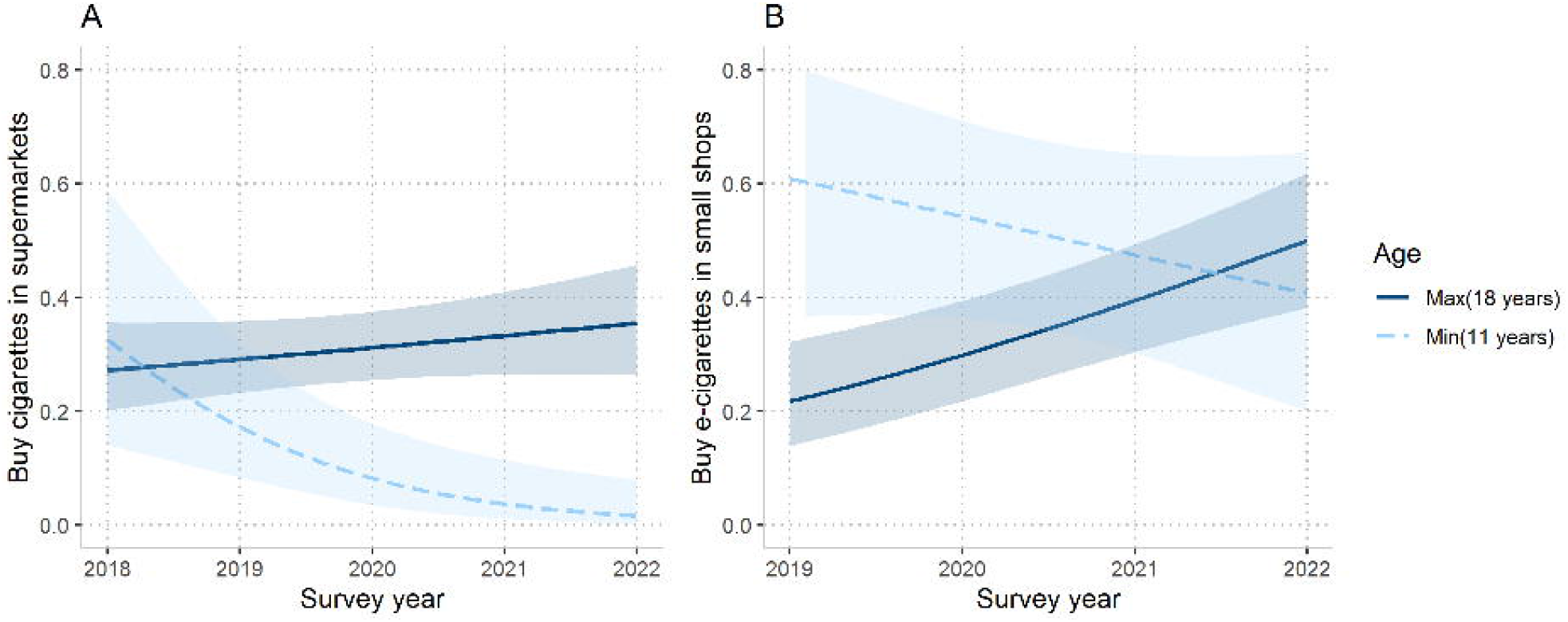
Interaction between age and survey year in logistic regression exploring sources of nicotine products in UK adolescents. A – Interaction between age and survey year for the likelihood of buying tobacco cigarettes from supermarkets (n=831). B – Interaction between age and survey year for the likelihood of buying e-cigarettes from small shops (n=608). *Note*: Only the predicted likelihoods for the minimum (11 years) and maximum (18 years) ages are shown for clarity.

#### Sources of e-cigarettes among current e-cigarette users

Overall, 12.3% of participants who used e-cigarettes bought them from supermarkets, 42.8% from small shops, 16.8% online and 55.3% acquired them from other sources (**Table 4**). In the adjusted logistic regression, there was some evidence that sources of e-cigarettes changed over the survey years. In 2022, participants were twice as likely to source e-cigarettes from small shops (OR 2.02; 95% CI 1.24, 3.29) and 68% less likely to source them from online (OR 0.32; 95% CI 0.17, 0.58), when compared to 2019. Younger e-cigarette users (16-17 and 14-15 years) were less likely to get their e-cigarettes from supermarkets and more likely to acquire them from other sources than 18-year-olds. Participants living in a household with someone who used e-cigarettes were more likely to source their e-cigarettes from supermarkets and small shops than those without an e-cigarette user in the household.

**Table 4.**
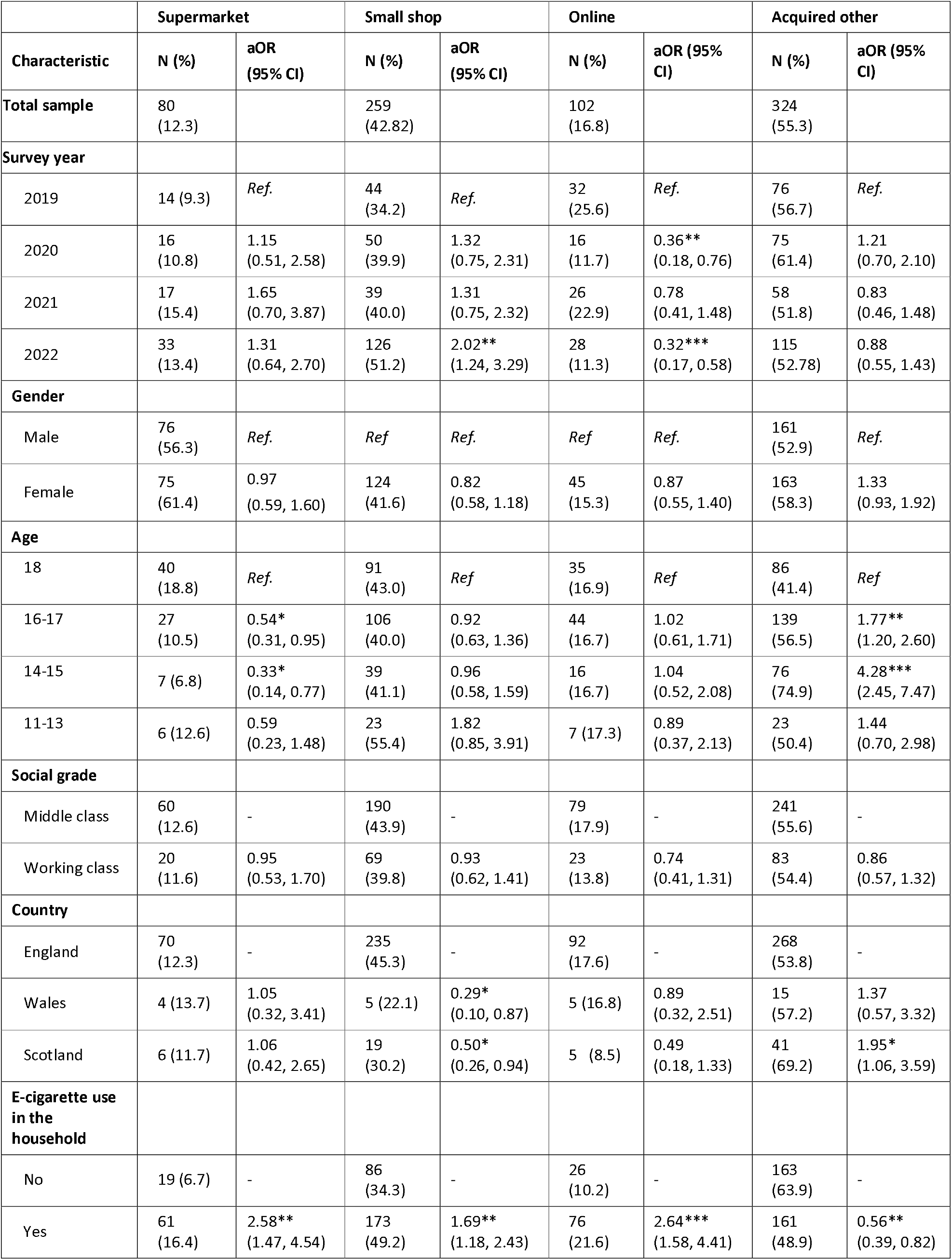
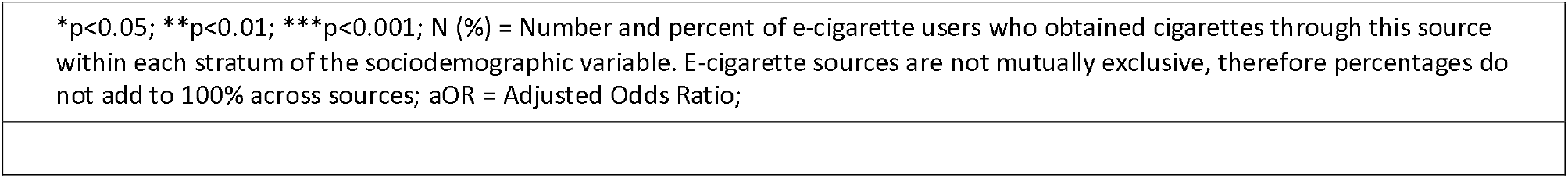
Adjusted logistic regression for likelihood of acquiring e-cigarettes from different sources in UK adolescents (n=608)

|  | Supermarket |  | Small shop |  | Online |  | Acquired other |  |
| --- | --- | --- | --- | --- | --- | --- | --- | --- |
| Characteristic | N (%) | aOR (95% CI) | N (%) | aOR (95% CI) | N (%) | aOR (95% CI) | N (%) | aOR (95% CI) |
| **Total sample** | 80 (12.3) |  | 259 (42.82) |  | 102 (16.8) |  | 324 (55.3) |  |
| **Survey year** |  |  |  |  |  |  |  |  |
| 2019 | 14 (9.3) | *Ref.* | 44 (34.2) | *Ref.* | 32 (25.6) | *Ref.* | 76 (56.7) | *Ref.* |
| 2020 | 16 (10.8) | 1.15 (0.51, 2.58) | 50 (39.9) | 1.32 (0.75, 2.31) | 16 (11.7) | 0.36** (0.18, 0.76) | 75 (61.4) | 1.21 (0.70, 2.10) |
| 2021 | 17 (15.4) | 1.65 (0.70, 3.87) | 39 (40.0) | 1.31 (0.75, 2.32) | 26 (22.9) | 0.78 (0.41, 1.48) | 58 (51.8) | 0.83 (0.46, 1.48) |
| 2022 | 33 (13.4) | 1.31 (0.64, 2.70) | 126 (51.2) | 2.02** (1.24, 3.29) | 28 (11.3) | 0.32*** (0.17, 0.58) | 115 (52.78) | 0.88 (0.55, 1.43) |
| **Gender** |  |  |  |  |  |  |  |  |
| Male | 76 (56.3) | *Ref.* | *Ref.* | *Ref.* | *Ref.* | *Ref.* | 161 (52.9) | *Ref.* |
| Female | 75 (61.4) | 0.97 (0.59, 1.60) | 124 (41.6) | 0.82 (0.58, 1.18) | 45 (15.3) | 0.87 (0.55, 1.40) | 163 (58.3) | 1.33 (0.93, 1.92) |
| **Age** |  |  |  |  |  |  |  |  |
| 18 | 40 (18.8) | *Ref.* | 91 (43.0) | *Ref.* | 35 (16.9) | *Ref.* | 86 (41.4) | *Ref.* |
| 16-17 | 27 (10.5) | 0.54* (0.31, 0.95) | 106 (40.0) | 0.92 (0.63, 1.36) | 44 (16.7) | 1.02 (0.61, 1.71) | 139 (56.5) | 1.77** (1.20, 2.60) |
| 14-15 | 7 (6.8) | 0.33* (0.14, 0.77) | 39 (41.1) | 0.96 (0.58, 1.59) | 16 (16.7) | 1.04 (0.52, 2.08) | 76 (74.9) | 4.28*** (2.45, 7.47) |
| 11-13 | 6 (12.6) | 0.59 (0.23, 1.48) | 23 (55.4) | 1.82 (0.85, 3.91) | 7 (17.3) | 0.89 (0.37, 2.13) | 23 (50.4) | 1.44 (0.70, 2.98) |
| **Social grade** |  |  |  |  |  |  |  |  |
| Middle class | 60 (12.6) | - | 190 (43.9) | - | 79 (17.9) | - | 241 (55.6) | - |
| Working class | 20 (11.6) | 0.95 (0.53, 1.70) | 69 (39.8) | 0.93 (0.62, 1.41) | 23 (13.8) | 0.74 (0.41, 1.31) | 83 (54.4) | 0.86 (0.57, 1.32) |
| **Country** |  |  |  |  |  |  |  |  |
| England | 70 (12.3) | - | 235 (45.3) | - | 92 (17.6) | - | 268 (53.8) | - |
| Wales | 4 (13.7) | 1.05 (0.32, 3.41) | 5 (22.1) | 0.29* (0.10, 0.87) | 5 (16.8) | 0.89 (0.32, 2.51) | 15 (57.2) | 1.37 (0.57, 3.32) |
| Scotland | 6 (11.7) | 1.06 (0.42, 2.65) | 19 (30.2) | 0.50* (0.26, 0.94) | 5 (8.5) | 0.49 (0.18, 1.33) | 41 (69.2) | 1.95* (1.06, 3.59) |
| **E-cigarette use in the household** |  |  |  |  |  |  |  |  |
| No | 19 (6.7) | - | 86 (34.3) | - | 26 (10.2) | - | 163 (63.9) | - |
| Yes | 61 (16.4) | 2.58** (1.47, 4.54) | 173 (49.2) | 1.69** (1.18, 2.43) | 76 (21.6) | 2.64*** (1.58, 4.41) | 161 (48.9) | 0.56** (0.39, 0.82) |

\*p<0.05; \*\*p<0.01; \*\*\*p<0.001; N (%) = Number and percent of e-cigarette users who obtained cigarettes through this source within each stratum of the sociodemographic variable. E-cigarette sources are not mutually exclusive, therefore percentages do not add to 100% across sources; aOR = Adjusted Odds Ratio;

Interactions between age and survey year were tested for all regression models and were found to be significant for buying e-cigarettes from small shops (**Figure 1.B**). In the younger participants, there was a decreasing trend in the odds of buying cigarettes from small shops between 2018-2022, but the confidence intervals overlapped. While there was an increase in likelihood for the older users buying e-cigarettes from small shops, from around 20% in 2019 to 50% in 2022.

##### Sensitivity analysis

To test the results were robust to possible misclassification bias, the variables for noticing e-cigarettes and tobacco cigarettes in supermarket and small shops were recoded, whereby not noticing additionally included the ‘hardly ever’ response. The proportion of participants who noticed both e-cigarettes and tobacco cigarettes were lower in the sensitivity analysis (**Supplementary Table 2**). For example, in the main analysis 64% of participants noticed cigarettes in supermarkets while 31% noticed them in the sensitivity analysis. Despite this change, the odds ratios in the main and sensitivity analyses were similar and the overall conclusions remained. For example, the lower likelihood of noticing tobacco cigarettes in supermarkets and small shops in 2022 compared to 2018 remained, as did the higher likelihood of noticing e-cigarettes on sale in supermarkets over time.

## DISCUSSION

In this study of adolescents in Great Britain, we found that the likelihood of seeing e-cigarettes on display in shops increased over time while the likelihood of seeing tobacco cigarettes on display decreased. There was not strong evidence that sources of tobacco cigarettes changed over time, but there was a higher likelihood that e-cigarette users obtained their e-cigarettes from small shops in 2022 compared to 2019. Younger adolescents were more likely to acquire their nicotine products from other sources and less likely to get them from supermarkets than those aged 18 years.

We found evidence that noticing tobacco cigarettes on sale fell over time but noticing e-cigarettes on display rose over time. This finding is reflective the current policy landscape in the UK whereby only tobacco cigarettes are covered by point of sale display bans. Exposure to e-cigarette displays have been cross-sectionally associated to e-cigarette use in Scottish adolescents [21], therefore a rise in noticing e-cigarette displays could impact e-cigarette uptake among children. However, e-cigarette use also rose during this time[18], therefore it is difficult to determine whether this is due to reverse causation. The rise in noticing e-cigarettes for sale was apparent for supermarkets but not small shops; small shops had consistently high levels of participants noticing e-cigarettes there. This is potentially reflective of the documented increased involvement of the traditional tobacco industry in the e-cigarette market.

Nicotine-product users aged 14-17 years were significantly less likely to buy their nicotine-product in supermarkets compared to users aged 18 years. Furthermore, it appears that the younger the user, the less likely they were to source their tobacco cigarettes from supermarkets over time, compared to older cigarette users. This suggests that minimum-age-restriction on purchasing tobacco did affect direct purchases from supermarkets, in line with previous findings [11]. Nonetheless, the fact that sources of tobacco cigarettes show no difference over time suggests that age of sale laws are currently underenforced, and support calls for the UK to raise this to 21 years. Additionally, introducing and enforcing “Challenge 25” measures to verify the age of potential customers would further address the consistent issue of availability of tobacco to children. For purchases of e-cigarettes from small shops there was no difference by age. This finding is consistent with the literature which has also shown underage users are able to purchases e-cigarettes. A study of the minimum age-restriction policy for e-cigarettes in Scotland suggested that in the first year the likelihood of children purchasing e-cigarettes was not affected by introduction of an age restriction[22]. Furthermore, an international study of tobacco use suggested that a similar number of over- and under-age individuals purchased their e-cigarettes from vape stores[23].

### Strengths and limitations

This study used a large representative sample of adolescents in Great Britain. The use of recent data (2018-2022) has permitted us to examine and account for the rising use of e-cigarettes in recent years, enhancing policy relevance. The dataset had detailed data on nicotine product use enabling us to conduct a detailed analysis on noticing and acquiring of these products, for the first time comparing the sources between countries, across years and by age.

There are some limitations to note. There was some missingness for questions regarding the display of nicotine products, consequently, the study lost power and the representativeness of the analysis may have been affected. Furthermore, due to low levels of tobacco and e-cigarettes use, the sample sizes for some of the nicotine product sources were low, but comparable to other nicotine product use surveys[19]. Participants were able to state multiple sources for their nicotine products, the binary variables used were not mutually exclusive. Therefore, this study cannot comment on the relationship between different sources. Participants also used their own judgement in determining what was a small shop and what was a supermarket, which may have introduced some misclassification. The dataset was limited in detailed collection of sociodemographic data and as such has missing information on ethnicity. Finally, it must be recognised that the COVID-19 pandemic might have affected the results. The 2020 data were collected in March 2020 and the 2021 data in March-April 2021. During this time, access to small shops and supermarkets were not restricted, however adolescents’ social lives may well have been impacted by the pandemic.

### Conclusions

This study on the awareness of nicotine-product display and sources of nicotine-products among adolescents in Great Britain suggests that measures to limit access to tobacco and e-cigarettes, such as age restrictions are not being adequately enforced. Policymakers need to be aware that additional policy approaches and enforcement will be required to effectively restrict awareness of and access to nicotine products for youth to blunt the trend for increased use in the age group.

## Data Availability

All data from this study was collected by YouGov and ASH

## Contributor statement

Study conception was by AL. Design was by AL and JP. JP performed the analyses and wrote the first draft. All other authors contributed to the development of the draft, interpretation of the results and approved the final version for submission.

