## Appendix for "Changing awareness and sources of tobacco and e-cigarettes among children in Great Britain"

| Four binary variables | Multi-categorical options | Tobacco cigarette – open ended responses | **E-cigarette – open ended responses** |
| --- | --- | --- | --- |
| Bought: supermarket | I buy them from a supermarket |  |  |
| Bought: small shop | I buy them from a newsagent, tobacconist, or a sweet shop |  | “Vape shops”  “Vape shops”  “Vape store”  “Vape shop” (3) |
|  | I buy them from a petrol station or garage shop |  |  |
| Bought: online  (e-cigs only) | I buy them through the Internet | N/A | “online” (2)  “On-line”  “Online from a vape store”  “Amazon” |
| Acquired other | I buy them from a machine | “Buy them abroad since they have been over 7 pound and got rid go 10 packs. As a light smoker It makes more sense as I never finish a pack”  “My friends go and buy them for me (those over 18)”  “Get big sister to go to shop for me”  “Older friends buy them for me” | “Someone else buys them from vape shop in town” |
|  | I buy them from some other type of shop |  |  |
|  | I buy them from street markets |  |  |
|  | I buy them from friends or relatives |  |  |
|  | I buy them from someone at school (not including friends) |  |  |
|  | I buy them from someone else |  |  |
|  | Friends give them to me | “Share with flatmates”  “Friends and strangers in smoking areas” | “I borrow my friends occasionally for a couple of puffs”  “I just use my friend's, who gets them from friends - both buying and being given them”  “Just lying round the house”  “Borrow from a friend”  “I only tend to smoke at parties” |
|  | My brother or sister gives them to me |  |  |
|  | My mother or father gives them to me |  |  |
|  | Someone else gives them to me |  |  |
|  | I take them | “Serbia”  “Rather illegally”  “I find them”  “I steal them from others” | “I don't buy them” |

SUPPLEMENTARY FILES

Supplementary Table 1 – Categorisation of responses to questions on sources of e-cigarettes and tobacco cigarettes

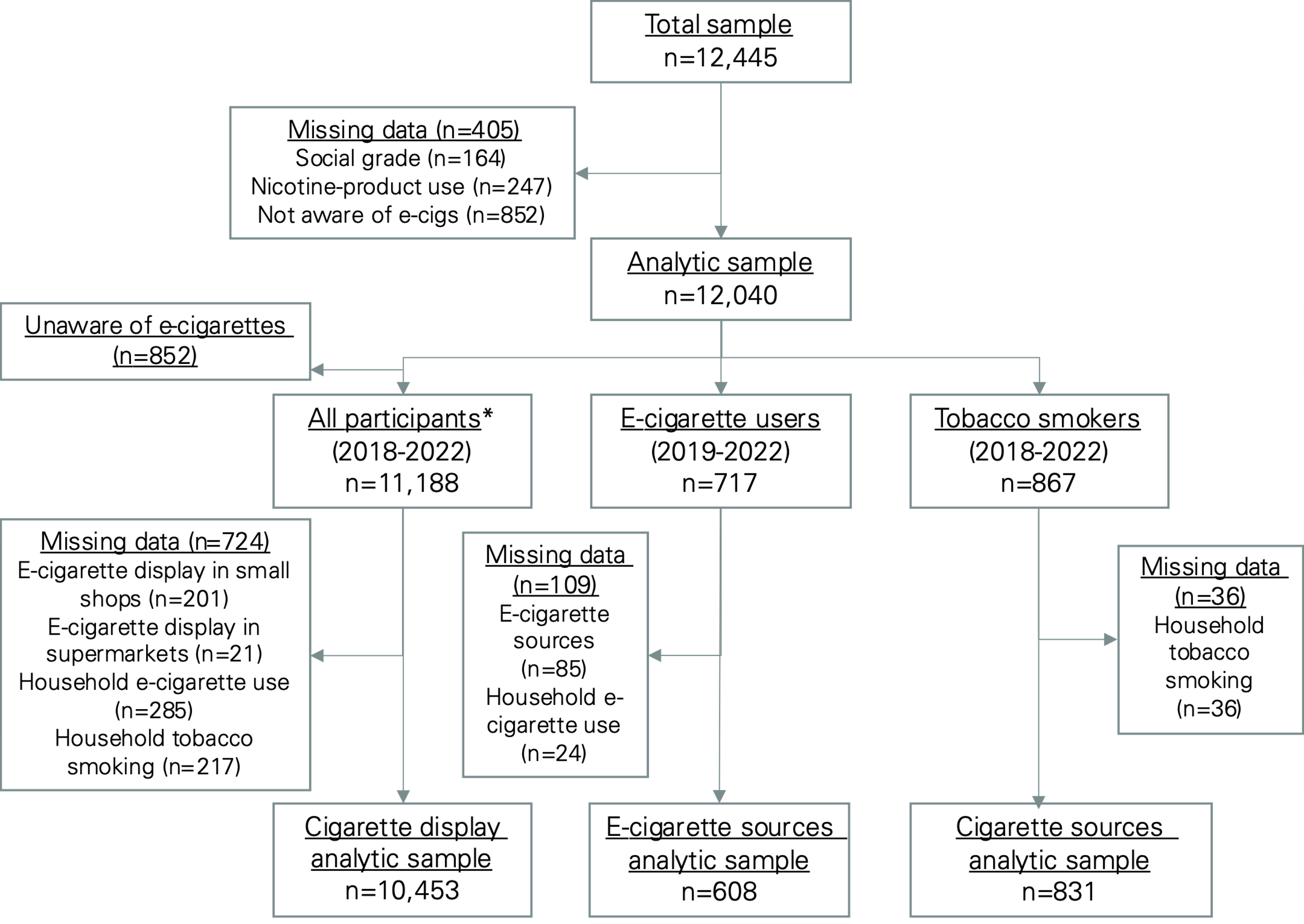

Supplementary Figure 1 – Flow diagram of sample exclusions in the analyses.

* All participants refers to all the participants who stated they were aware of e-cigarettes.

Supplementary Table 2 – Adjusted logistic regression showing the likelihood of noticing e-cigarettes and tobacco cigarettes on sale in supermarkets and small shops (n=10,453), with noticing recoded to include ‘every time’, ‘most times’ and ‘sometimes’ compared against ‘hardly ever’ and ‘never’

|  | Model 1: Tobacco cigarettes | | | | Model 2: E-cigarettes | | | |
| --- | --- | --- | --- | --- | --- | --- | --- | --- |
|  | Supermarket | | Small shop | | Supermarket | | Small shop | |
| Characteristic | N  (%) | aOR  (95% CI) | N  (%) | aOR  (95% CI) | N  (%) | aOR  (95% CI) | N  (%) | aOR  (95% CI) |
| Total Sample | 3453  (31.05) |  | 5453 (49.55) |  | 3683 (33.76) |  | 5100 (46.86) |  |
| **Survey year** |  |  |  |  |  |  |  |  |
| 2018 | 583 (32.97) | *Ref.* | 1025 (56.96) | *Ref.* | 422 (23.73) | *Ref.* | 779 (43.61) | *Ref.* |
| 2019 | 803 (33.35) | 1.04 (0.89, 1.21) | 1175 (49.44) | 0.73*** (0.63, 0.84) | 808 (34.46) | 1.78*** 1.53, 2.08) | 1025 (43.84) | 1.02 (0.89, 1.17) |
| 2020 | 786 (33.17) | 1.04 (0.90, 1.20) | 1171 (49.97) | 0.75*** (0.66, 0.87) | 917 (39.57) | 2.26*** 1.94, 2.63) | 1157 (50.26) | 1.35*** (1.18, 1.55) |
| 2021 | 658 (28.5) | 0.82** (0.70, 0.95) | 1021 (45.07) | 0.61*** (0.53, 0.70) | 729 (32.66) | 1.67*** 1.43, 1.96) | 978 (43.67) | 1.04 (0.90, 1.19) |
| 2022 | 623 (27.38) | 0.78** (0.67, 0.91) | 1061 (47.57) | 0.70*** (0.61, 0.81) | 807 (36.36) | 1.91*** 1.63, 2.23) | 1161 (52.28) | 1.46*** (1.27, 1.68) |
| Gender |  |  |  |  |  |  |  |  |
| Male | 1573 (29.46) | *Ref.* | 2567 (48.21) | *Ref.* | 1746 (33.05) | *Ref.* | 2454 (46.32) | *Ref.* |
| Female | 1880 (32.7) | 1.21*** (1.10, 1.32) | 2886 (50.94) | 1.15** (1.05, 1.25) | 1937 (34.5) | 1.08 0.99, 1.19) | 2646 (47.42) | 1.06 (0.98, 1.16) |
| Age |  |  |  |  |  |  |  |  |
| 18 | 915 (46.45) | *Ref.* | 1364 (69) | *Ref.* | 849 (43.5) | *Ref.* | 1156 (58.76) | *Ref.* |
| 16-17 | 1342 (47.26) | 1.07 (0.94, 1.21) | 1918 (67.49) | 0.96 (0.85, 1.09) | 1272 (43.92) | 1.10 0.97, 1.25) | 1699 (58.86) | 1.07 (0.94, 1.21) |
| 14-15 | 561 (22.14) | 0.35*** (0.30, 0.40) | 1045 (40.75) | 0.33*** (0.29, 0.37) | 789 (30.74) | 0.64*** 0.56, 0.73) | 1128 (43.87) | 0.60*** (0.53, 0.68) |
| 11-13 | 635 (21.35) | 0.34*** (0.29, 0.38) | 1126 (37.29) | 0.29*** (0.25, 0.33) | 773 (25.95) | 0.52*** 0.45, 0.59) | 1117 (37.08) | 0.46*** (0.41, 0.53) |
| Social grade |  |  |  |  |  |  |  |  |
| ABC1  (higher) | 2545 (32.36) | *Ref.* | 3973 (50.93) | *Ref.* | 2591 (33.62) | *Ref.* | 3624 (46.78) | *Ref.* |
| C2DE  (lower) | 908 (27.9) | 0.80*** (0.72, 0.89) | 1480 (46.25) | 0.85*** (0.77, 0.93) | 1092 (34.09) | 1.00 0.91, 1.11) | 1476 (47.04) | 1.01 (0.91, 1.11) |
| Country |  |  |  |  |  |  |  |  |
| England | 3029 (31.49) | *Ref.* | 4806 (50.59) | *Ref.* | 3192 (33.93) | *Ref.* | 4431 (47.17) | *Ref.* |
| Wales | 151 (29.55) | 0.92 (0.75, 1.14) | 239 (46.17) | 0.97 (0.77, 1.21) | 173 (33.23) | 1.00 (0.81, 1.25) | 248 (48.58) | 1.09 (0.87, 1.38) |
| Scotland | 273 (27.12) | 0.69*** (0.60, 0.81) | 408 (40.38) | 0.65*** (0.56, 0.77) | 318 (32.25) | 0.77*** (0.66, 0.89) | 421 (42.37) | 0.71*** (0.61, 0.84) |
| Current nicotine-product use status |  |  |  |  |  |  |  |  |
| Non-user | 2924 (29.27) | *Ref.* | 4702 (47.65) | *Ref.* | 3008 (30.95) | *Ref.* | 4277 (44.1) | *Ref.* |
| E-cigarette only | 161 (48.33) | 1.51** (1.16, 1.97) | 230 (68.63) | 1.65*** (1.25, 2.16) | 219 (65.19) | 2.63*** 2.03, 3.41) | 265 (79.08) | 3.06*** (2.29, 4.10) |
| Tobacco cigarettes only | 189 (45.75) | 1.17 (0.92, 1.47) | 291 (69.56) | 1.45** (1.13, 1.85) | 225 (54.96) | 2.12*** 1.69, 2.66) | 280 (66.03) | 1.82*** (1.45, 2.29) |
| Dual-user | 179 (55.29) | 1.76*** (1.35, 2.30) | 230 (69.85) | 1.57** (1.19, 2.07) | 231 (70.5) | 3.03*** 2.32, 3.95) | 278 (83.14) | 3.78*** (2.77, 5.17) |
| E-cigarette use in the household |  |  |  |  |  |  |  |  |
| No | 2762 (29.84) | *Ref.* | 4437 (48.83) | *Ref.* | 2748 (30.45) | *Ref.* | 3959 (44.11) | *Ref.* |
| Yes | 691 (36.63) | 2.12*** (1.80, 2.51) | 1016 (52.9) | 1.60*** (1.35, 1.90) | 935 (49.12) | 1.56*** 1.32, 1.83) | 1141 (59.62) | 1.47*** (1.24, 1.73) |
| Tobacco smoking in the household |  |  |  |  |  |  |  |  |
| No | 3042 (29.62) | *Ref.* | 4928 (48.56) | *Ref.* | 3262 (32.43) | *Ref.* | 4582 (45.65) | *Ref.* |
| Yes | 411 (47.7) | 1.20** (1.05, 1.36) | 525 (61.07) | 1.08 (0.96, 1.22) | 421 (49.31) | 1.86*** 1.64, 2.10) | 518 (60.92) | 1.56*** (1.39, 1.76) |
| N (%) = Number and percent of participants who noticed e-cigarettes or tobacco cigarettes on display within each stratum of the sociodemographic variables; aOR = adjusted Odds Ratio; 95% CI = 95% confidence interval; *p<0.05; **p<0.01; ***p<0.001; | | | | | | | | |
